# A Descriptive Study of a Clinical Pharmacist’s Role in the Outpatient Pulmonary Hypertension Clinic in the United States (CliPR-PH)

**DOI:** 10.1101/2023.06.22.23291725

**Authors:** Vienica Funtanilla, Dylan Lee, Eric Kim, Crystal Zhou

**Affiliations:** University of California, San Francisco Department of Clinical Pharmacy and Janssen Scientific Affairs; University of California, San Francisco School of Pharmacy; University of California, San Francisco School of Pharmacy and Department of Clinical Pharmacy

**Keywords:** collaborative practice, selexipag, titration, managing side effects

## Abstract

**BACKGROUND:** Pulmonary arterial hypertension (PAH) is a rare disease affecting the heart and lungs. Median survival was 2.8 years historically, but prognosis improved with the advances of PAH therapy. It is currently not standard of care for clinical pharmacists to be a part of the interdisciplinary care team in outpatient PH clinics to help manage patients’ drug regimens, and there is limited literature that has explored the impact of clinical pharmacists on patient-related processes and outcomes. The goal of the CliPR-PH study is to characterize the activities of the clinical pharmacist as a member of the interdisciplinary team.

**METHODS:** CliPR-PH was a retrospective, descriptive, single-center cohort study of patients ≥ 18 years, diagnosed with pulmonary hypertension, and managed by a clinical pharmacist practicing under a collaborative practice agreement between January 1, 2018 and July 31, 2020. Patients were excluded from the study if they were not on any PAH medications at the time of pharmacist encounter.

**RESULTS:** Sixty patients were included in the analysis and 331 clinical pharmacist interventions were documented over the study period. Interventions were regarding selexipag [88 (26.6%)], sildenafil [74 (22.4%)], tadalafil [46 (13.9%)], prior authorization (PA) completions [127 (38.4%), PA troubleshooting [43 (13.0%)], and ‘Other’ [49 (14.8%)].

**CONCLUSION:** Clinical pharmacists can play an important role in closely monitoring patients during the medication titration phase and ensure prior authorizations are approved in a timely manner as part of the interdisciplinary team in the outpatient PH clinic setting using a collaborative practice model approach.

**Clinical Perspective:** What is new?

There are currently no studies in the United States that asses a clinical pharmacist’s role on the interdisciplinary team as part of a collaborative practice agreement in the outpatient pulmonary hypertension (PH) clinic.

What are the clinical implications?

Integrating a clinical pharmacist on the medical team in an outpatient PH clinic setting allows them to closely monitor patients who are titrating selexipag, manage adverse events (AEs) that occur during the titration phase, and ensure medications requiring prior authorizations are approved in a timely manner.

## INTRODUCTION

Pulmonary hypertension (PH) is a disease that affects the heart and lungs and is classified into 5 groups. The approved drugs for PH are indicated for World Health Organization (WHO) Group 1, pulmonary arterial hypertension (PAH).^1^ PAH is a rare disease with an estimated prevalence of 48 to 55 cases per million adults based on registry data from economically developed countries.^1^ It is characterized by elevated pulmonary arterial pressure and assessed via right heart catheterization. Before targeted PAH therapy was available, the prognosis for PH was poor with a median survival of 2.8 years and estimated 1-year, 3-year, and 5-year survival rates of 68%, 48%, and 34%, respectively, according to the NIH registry. The prognosis improved after advances in PAH therapy in the 1990s, with 1-year, 3-year, 5-year, and 7-year survival rates of 85%, 68%, 57%, and 49%, respectively, according to patients enrolled in the REVEAL registry between 2006 and 2009.^2^

Treatment regimens for PAH tend to be complex, in part because there are five classes of medications that are approved to treat PAH and they are frequently used in combination.^3^ In patients without cardiopulmonary comorbidities, the 2022 ESC/ERS Guidelines for the treatment of PH recommend initial oral combination therapy in patients who are at low or intermediate risk of death at 1 year and initial triple therapy with a parenteral agent in patients at high risk of death at 1 year. ^1^ In addition to targeted therapy, supportive therapy such as oral anticoagulation, diuretics, digoxin, and management of anemia may also be warranted in patients who experience other cardiovascular complications due to PAH. ^1, 4^

Due to the complexity of medication treatment in PAH patients, clinical pharmacists can play a vital role as part of the interdisciplinary team. The different roles that clinical pharmacists can play in the treatment of patients with PAH include formulary management, medication safety such as the creation of medication order sets to lower chances of medication error, regulatory compliance, patient and care provider support, medication access, side effect management, drug interaction assessment, and transitions of care support.^4, 5^ It is currently not standard of care for clinical pharmacists to be a part of the interdisciplinary care team in outpatient PH clinics to help manage patients’ drug regimens, and there is limited literature that has explored the impact of clinical pharmacists on patient-related processes and outcomes. The goal of the CliPR-PH study is to further characterize the activities of the clinical pharmacist as a member of the interdisciplinary team.

### Study design

CliPR-PH was a retrospective, descriptive, single-center cohort study of patients ≥ 18 years, diagnosed with PH, and managed by an interdisciplinary team that included a clinical pharmacist between January 1, 2018 and July 31, 2020. The first date of encounter by the clinical pharmacist was identified as the index date.

### Methods

Patients ≥ 18 years of age with an established diagnosis of PH at the date of the first pharmacist’s encounter, who had at least one clinic visit between January 1, 2018, to July 31, 2020, were included in the study (Figure 1). To identify the study cohort, a list of patients was obtained from the University of California San Francisco (UCSF) Medical Center Information Technology Services for patients seen between this time period, with PH or PAH (ICD-9 codes: 416.8, 747.83, 416.0, 416.8; ICD-10 codes: I27.20, P29.30, I27.2, I27.0, I27.24, I27.29, I27.21, I27.23, I27.22), and who interacted with a clinical pharmacist at the UCSF outpatient PH center. An encrypted spreadsheet was created that censored patient identifier information (medical record number and name) to collect patient information and pharmacist interventions. Interventions that were collected included medication changes (dose increase, switching therapy, or discontinuation), prior authorizations, enrollment in patient assistance programs, and side effect management. Patients were excluded from the study if they were not on any PAH medications at the time of their pharmacist encounter. Chart review was conducted by DL, EK, and SC and VF and CZ verified for accuracy. Descriptive statistics were conducted using Microsoft Excel. This study was reviewed and approved by the WCG Institutional Review Board (IRB# 21-35283) as an exempt study under 45 CFR 46.104(d)(4).

**Figure 1.**
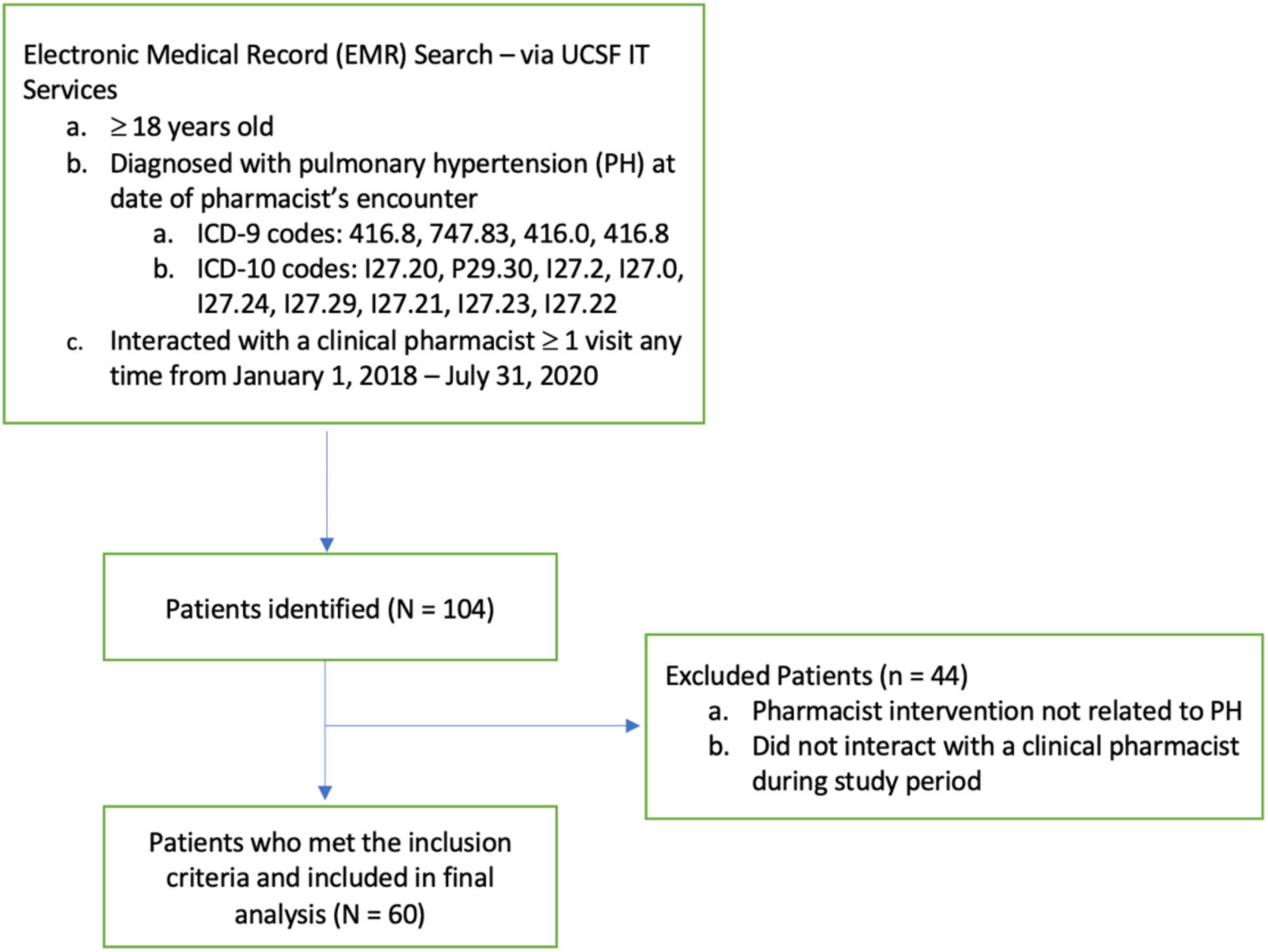
Inclusion/Exclusion Criteria.

## RESULTS

### Study population and characteristics

Sixty patients met the inclusion criteria and were included in the analysis. Among the 60 patients, there were 331 documented clinical pharmacist interventions over the study period. The study population at baseline consisted of patients with a mean age of 54 years, a higher number of females (60%), and predominantly WHO FC II and III (Table 1).

**Table 1.**
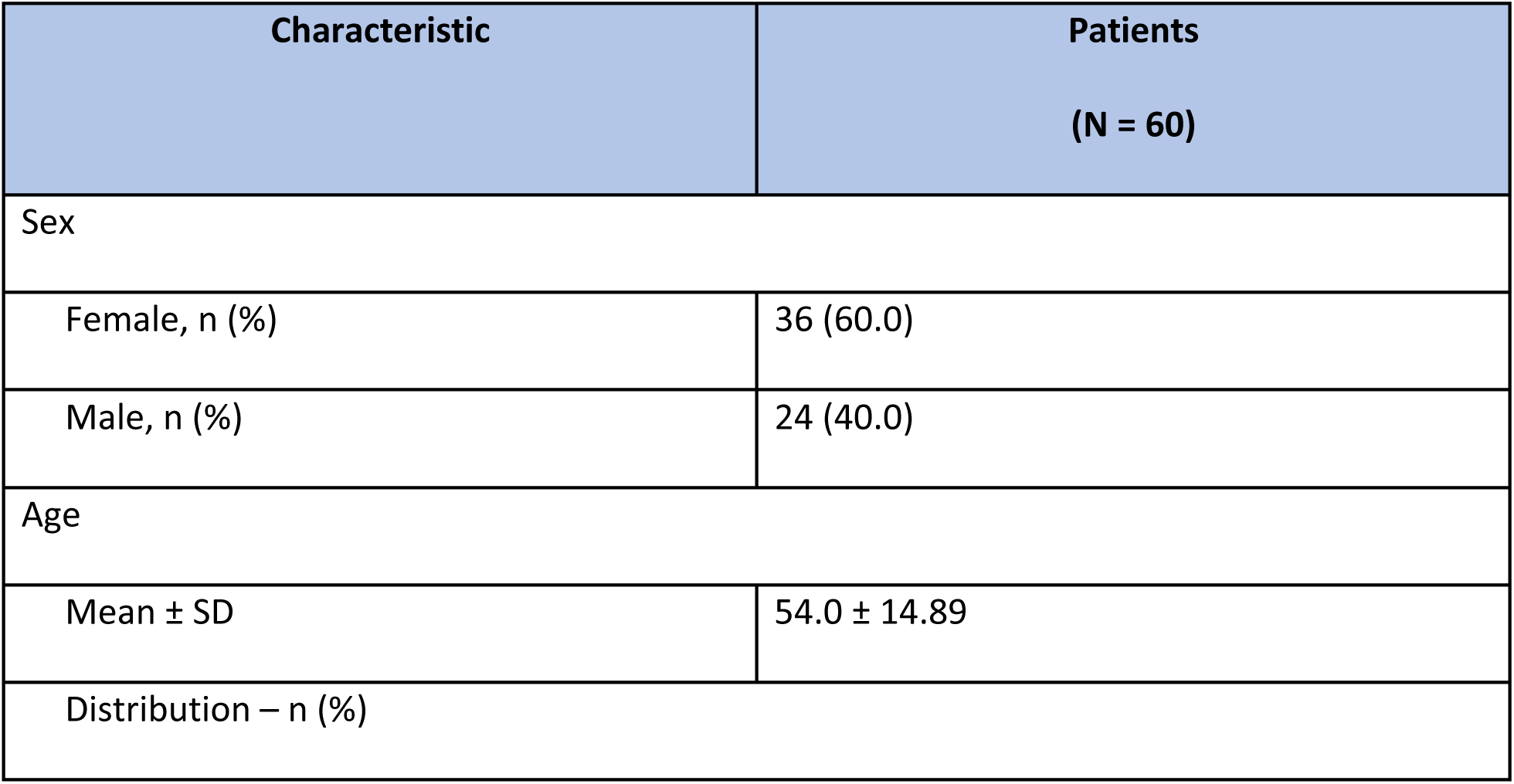
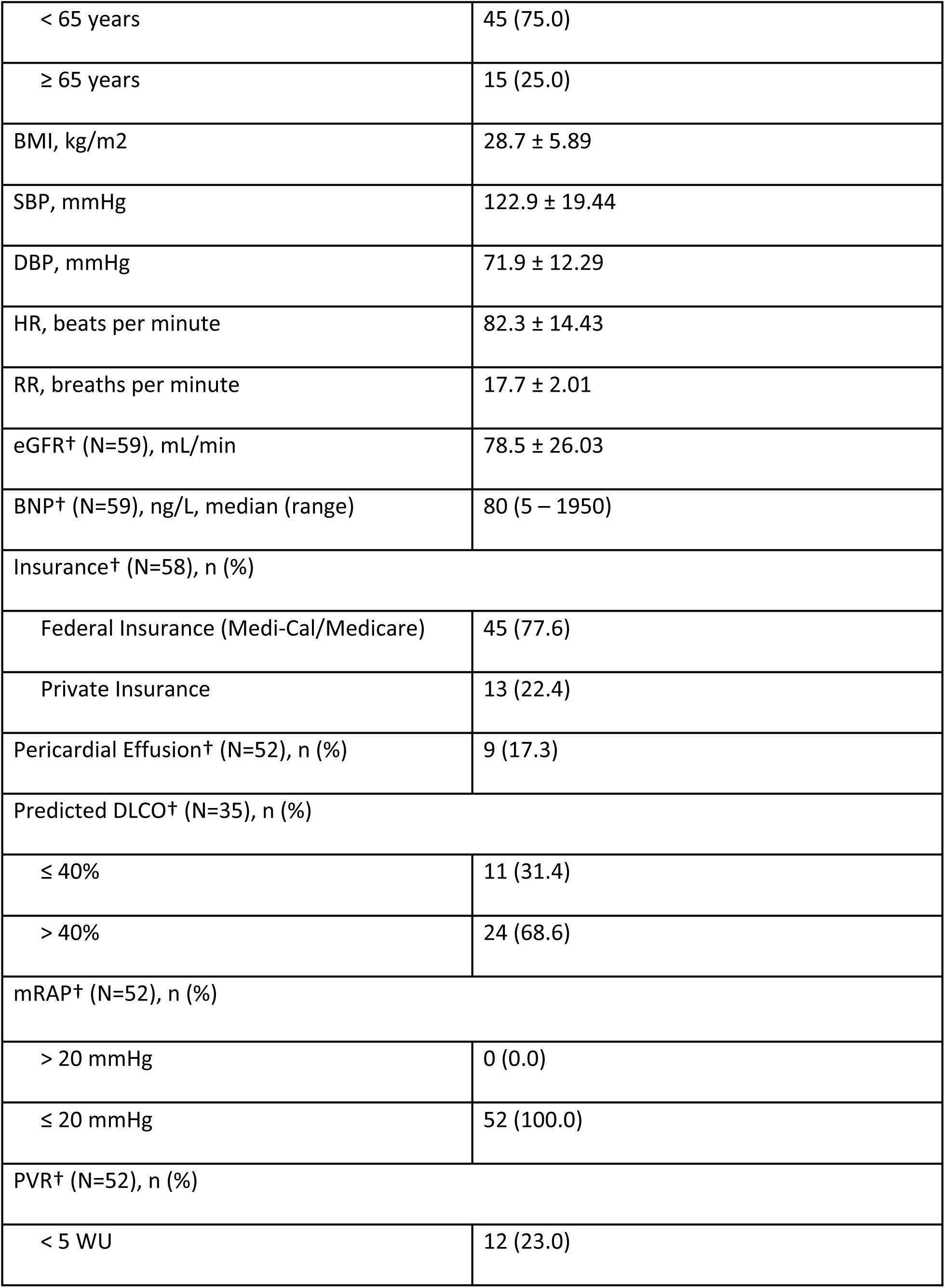
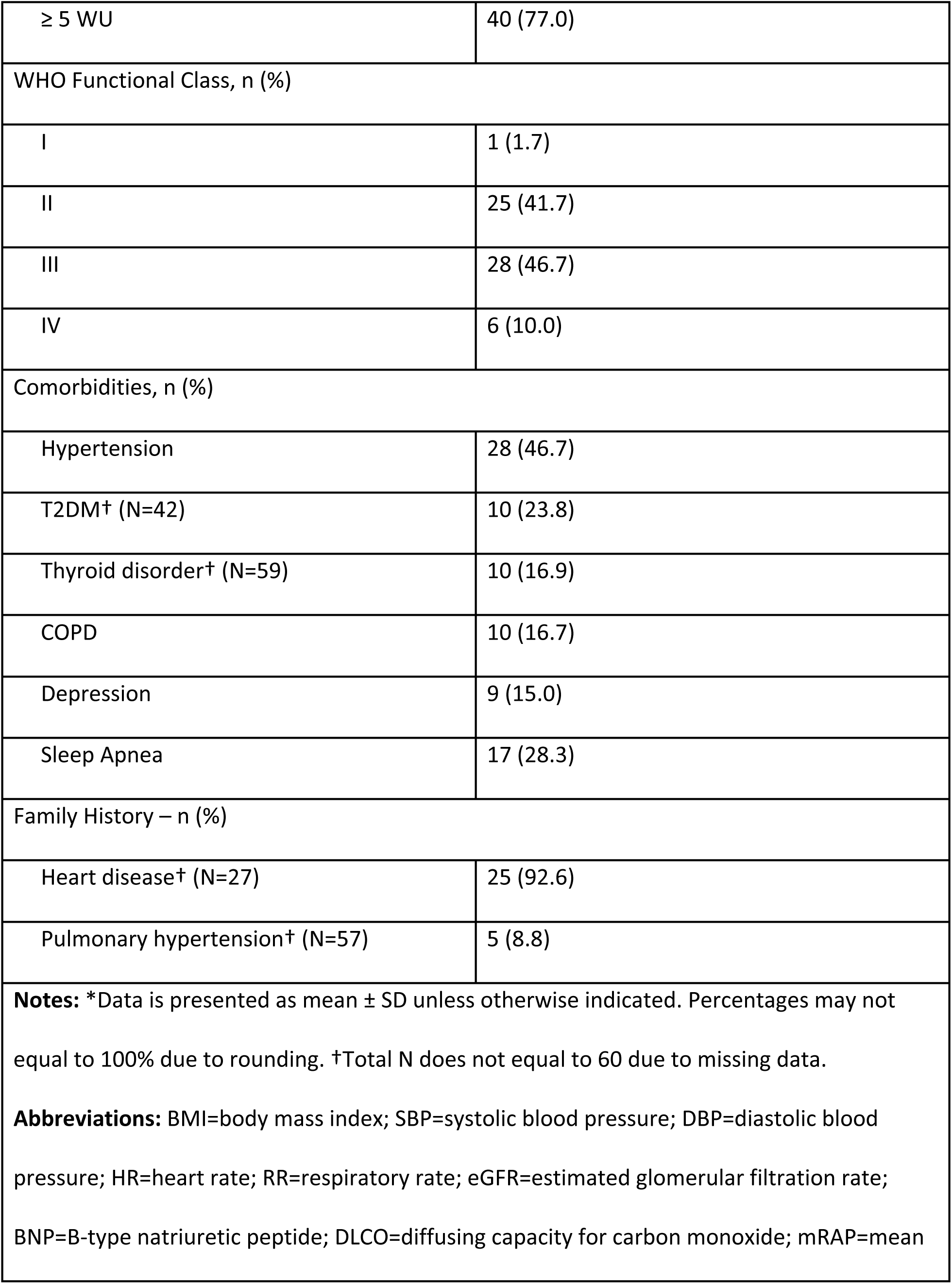
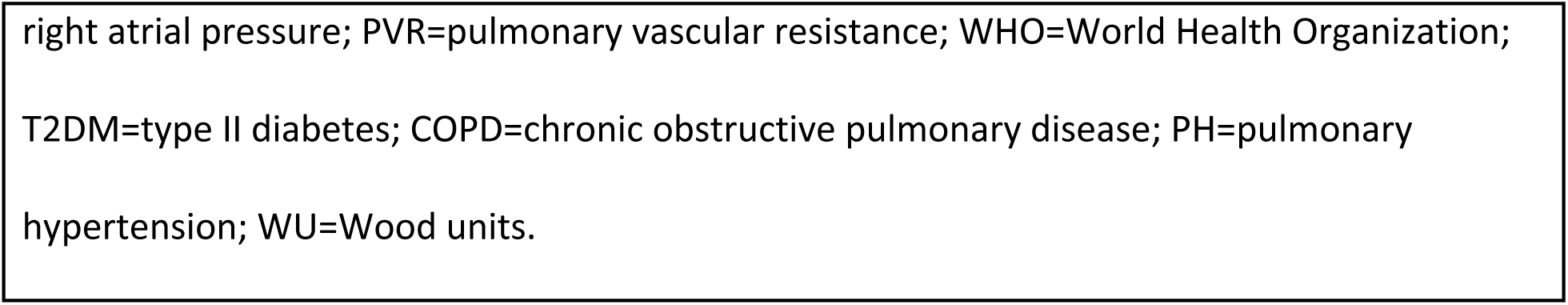
Baseline characteristics.

| Characteristic | Patients<br>(N = 60) |
| --- | --- |
| Sex |  |
| Female, n (%) | 36 (60.0) |
| Male, n (%) | 24 (40.0) |
| Age |  |
| Mean ± SD | 54.0 ± 14.89 |
| Distribution – n (%) |  |
| < 65 years | 45 (75.0) |
| ≥ 65 years | 15 (25.0) |
| BMI, kg/m<sup>2</sup> | 28.7 ± 5.89 |
| SBP, mmHg | 122.9 ± 19.44 |
| DBP, mmHg | 71.9 ± 12.29 |
| HR, beats per minute | 82.3 ± 14.43 |
| RR, breaths per minute | 17.7 ± 2.01 |
| eGFR<sup>†</sup> (N=59), mL/min | 78.5 ± 26.03 |
| BNP<sup>†</sup> (N=59), ng/L, median (range) | 80 (5 – 1950) |
| Insurance<sup>†</sup> (N=58), n (%) |  |
| Federal Insurance (Medi-Cal/Medicare) | 45 (77.6) |
| Private Insurance | 13 (22.4) |
| Pericardial Effusion<sup>†</sup> (N=52), n (%) | 9 (17.3) |
| Predicted DLCO<sup>†</sup> (N=35), n (%) |  |
| ≤ 40% | 11 (31.4) |
| > 40% | 24 (68.6) |
| mRAP<sup>†</sup> (N=52), n (%) |  |
| > 20 mmHg | 0 (0.0) |
| ≤ 20 mmHg | 52 (100.0) |
| PVR<sup>†</sup> (N=52), n (%) |  |
| < 5 WU | 12 (23.0) |
| ≥ 5 WU | 40 (77.0) |
| WHO Functional Class, n (%) |  |
| I | 1 (1.7) |
| II | 25 (41.7) |
| III | 28 (46.7) |
| IV | 6 (10.0) |
| Comorbidities, n (%) |  |
| Hypertension | 28 (46.7) |
| T2DM<sup>†</sup> (N=42) | 10 (23.8) |
| Thyroid disorder<sup>†</sup> (N=59) | 10 (16.9) |
| COPD | 10 (16.7) |
| Depression | 9 (15.0) |
| Sleep Apnea | 17 (28.3) |
| Family History – n (%) |  |
| Heart disease<sup>†</sup> (N=27) | 25 (92.6) |
| Pulmonary hypertension<sup>†</sup> (N=57) | 5 (8.8) |
| **Notes:** *Data is presented as mean ± SD unless otherwise indicated. Percentages may not equal to 100% due to rounding. <sup>†</sup>Total N does not equal to 60 due to missing data.<br>**Abbreviations:** BMI=body mass index; SBP=systolic blood pressure; DBP=diastolic blood pressure; HR=heart rate; RR=respiratory rate; eGFR=estimated glomerular filtration rate; BNP=B-type natriuretic peptide; DLCO=diffusing capacity for carbon monoxide; mRAP=mean |  |

right atrial pressure; PVR=pulmonary vascular resistance; WHO=World Health Organization; T2DM=type II diabetes; COPD=chronic obstructive pulmonary disease; PH=pulmonary hypertension; WU=Wood units.

### Pharmacist Interventions

The clinical pharmacist supported a diverse range of medications and medication issues. Out of 331 interventions, 88 (26.6%) interventions were regarding selexipag, 74 (22.4%) interventions were regarding sildenafil, and 46 (13.9%) interventions were regarding tadalafil [Figure 2]. One-hundred twenty-seven (38.4%) pharmacist interventions were prior authorization (PA) completions, 43 (13.0%) pharmacist interventions were PA troubleshooting, and 49 (14.8%) pharmacist interventions were ‘Other’, [Figure 3]. ‘Other’ consisted of a variety of pharmacist interventions including medication reconciliation, medication access troubleshooting not related to insurance (i.e., bubble packs), phone calls to patients for lab reminders and to pharmacies for medication approval verification, blood pressure check, and blood pressure machine troubleshooting.

**Figure 2.**
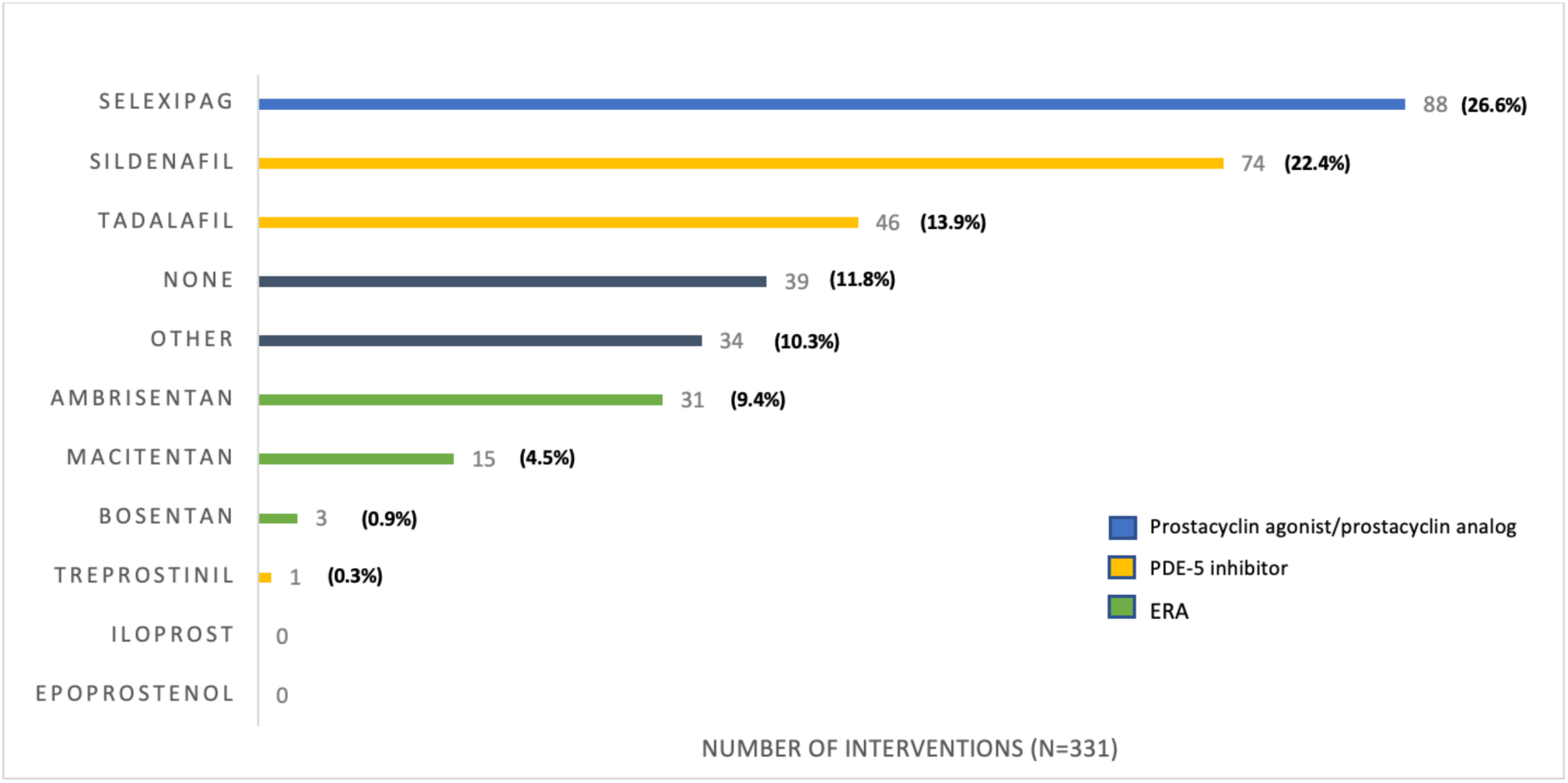
Medications involved in pharmacist interventions. **Notes:** None indicates that no medication was associated with the pharmacist intervention (i.e., Intervention was regarding medication reconciliation, lab reminders, etc.). Other indicates medication that were not PAH-targeted therapies. *Selexipag is a prostacyclin receptor agonist. Treprostinil, iloprost, and epoprostenol are prostacyclin analogs. **Abbreviations:** ERA=endothelin receptor antagonist; PDE-5=phosphodiesterase-5.

**Figure 3.**
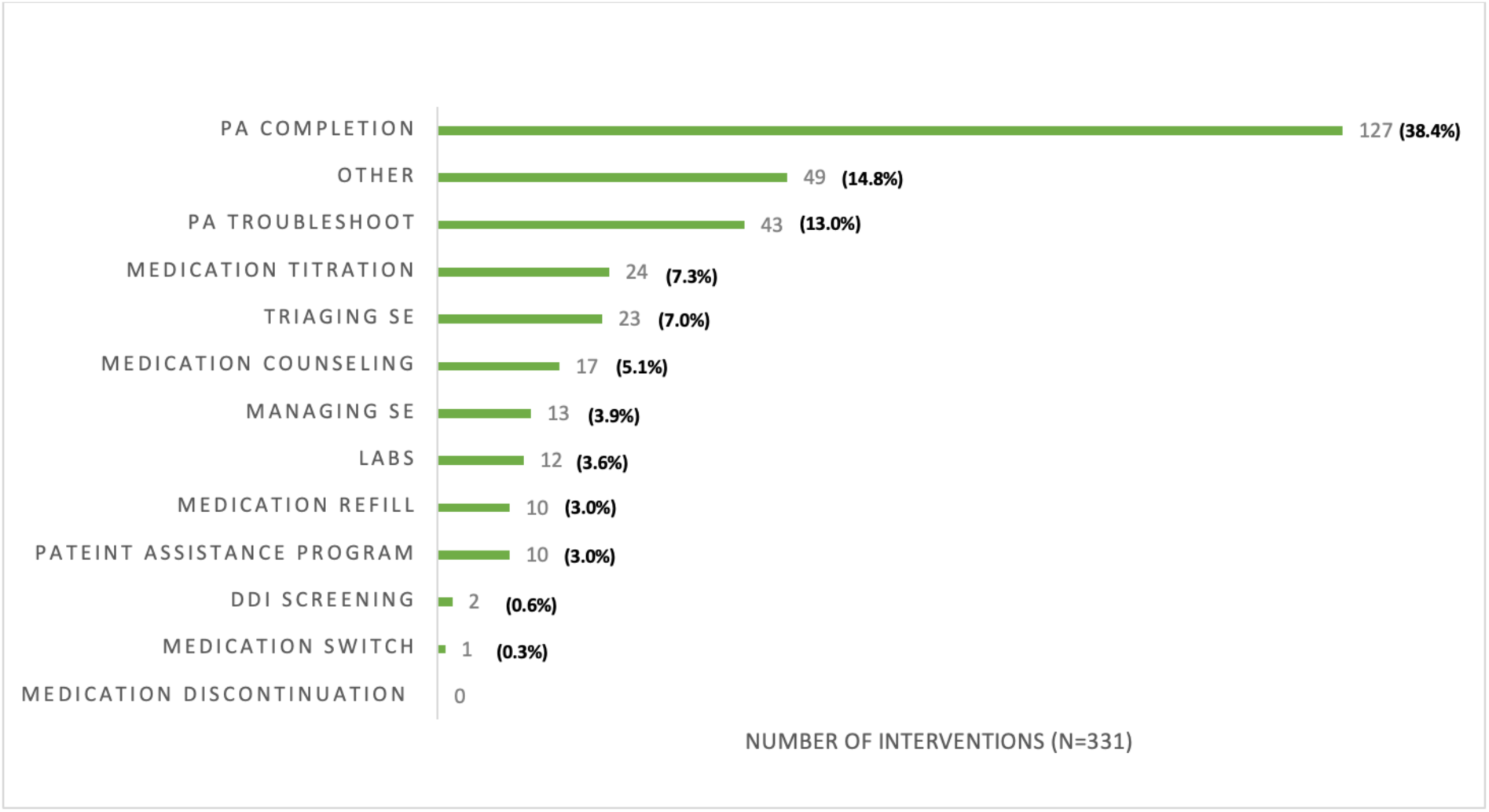
Pharmacist interventions. **Note:** “Other” interventions included medication reconciliation, medication access troubleshooting not related to insurance (i.e., bubble packs), call reminders to patients, calls to pharmacies and insurance companies to verify medication approvals, blood pressure check, and blood pressure machine troubleshooting. **Abbreviations:** DDI=drug-drug interaction; PA=prior authorization; SE=side effect.

## DISCUSSION

The CliPR-PH study is the first study that examines clinical pharmacist work through a collaborative practice agreement supporting PH patients in the United States. In France, Roustit et al conducted a prospective, multicenter randomized controlled study that assessed the impact of clinical pharmacists in the collaborative care of patients diagnosed with PH and were on initial PH-specific (i.e., endothelin receptor antagonist, phosphodiesterase-5 inhibitors, prostacyclin analogs, or a combination of these drugs) and non-specific (i.e., diuretics and anticoagulants) therapy between March 2010 and August 2012. Drug-related problems (DRP) were more frequently resolved in the collaborative care group with a clinical pharmacist versus the standard of care group (86.5% vs 66.7%, p=0.01). Resolution of DRPs also translated into reduced costs associated with DRP-related hospitalization. However, there was no demonstrated difference in adherence, satisfaction with medication, and health-related quality of life between groups. ^6^ Since the enrollment of patients between 2010 and 2012 in the Roustit et al study, new treatments in PH emerged, specifically selexipag’s approval in 2015. Selexipag is a prostacyclin receptor agonist with a starting dose of 200 mcg twice daily and titrated to the highest tolerated dose, up to 1600 mcg twice daily.^7^ With the approval of selexipag, the role of the clinical pharmacist has evolved. Selexipag requires dosing titration and clinical pharmacists have the expertise to closely monitor patients, perform dose titration strategies, and manage AEs during the titration phase. Selexipag is also recommended as an add-on therapy to the initial combination therapy per the 2022 ERS/ESC Guidelines, suggesting that patients on selexipag have more severe disease and are on more complex regimens.

Registry data shows that patients with PAH have a high chance of having a comorbid condition along with their PAH diagnosis. In the SPHERE registry of patients who initiated selexipag in the United States, 98.8% of 500 patients had at least one comorbidity and 55% were receiving combination therapy. Most patients at baseline were also WHO FC II (31%) and III (49.6%).^8^ In our study, the most common medication that was involved the pharmacist’s interventions was selexipag and 46.7% of patients at baseline were WHO FC III, suggesting that the population of patients encountered by the pharmacist had more severe disease and a more complex medication regimen. Additionally in UCSF’s collaborative practice agreement, patients are referred to the pharmacist by the physician and usually, patients requiring referrals are those with more severe disease and whose treatments are more complex. Patients with more complex medication regimens provide an opportunity for pharmacists to utilize their knowledge in multiple disease states and play a role in identifying drug interactions and minimizing polypharmacy.

In patients who are initiating oral selexipag, there is a role for clinical pharmacists in the outpatient setting to help with dose titration and manage AEs. Since the recommended starting dose of selexipag is 200 mcg twice daily and patients titrate usually at weekly intervals of 200 mcg twice daily to the highest tolerated dose up to 1600 mcg twice daily, it would take at least 7 weeks for a patient to reach the highest tolerated dose. The dose can also be reduced if patients are not able to tolerate the titrated dose, which can take longer to reach the highest tolerated dose.^7^ In a retrospective study conducted by Palevsky et al of specialty pharmacy data examining real-world dosing patterns of selexipag, 79% of 523 patients who had history of prior prostacylin use and 87.1% of 3,408 patients who have no history of prior prostacyclin use titrated slower than weekly intervals (>8 days/200 mcg twice daily).^9^ This suggests that titration of selexipag is personalized per patient and clinical pharmacists as part of the interdisciplinary medical team can closely monitor these patients to ensure they are titrating at an appropriate speed to reach their individual maintenance doses. In a real-world study conducted by Chang et al of 113 Korean patients with PAH who initiated selexipag and were followed up for 24 weeks, it was found that AEs occurred more often in the titration phase and that recovery rates were 65-77% with a median time to recovery of 15-70 days (range, 2-233 days). The median time to resolution of an AE was 15 days for nausea/vomiting and about 2 months for diarrhea, myalgia, and headache.^10^ As highlighted by this data, the AEs associated with selexipag occur more often in the titration phase and takes a few weeks to recover. Patients may not be willing to continue taking a medication with AEs especially if they are not aware of what to expect. Clinical pharmacists can closely monitor patients on selexipag as they titrate to their maintenance dose and help manage AEs.

In addition to closely monitoring patients during selexipag titration, the clinical pharmacist in our study played an essential role in improving medication access for patients through facilitating insurance approval. Pharmacists are knowledgeable with the insurance approval process and can use best practice for prior authorization approvals so that patients receive their medications in a timely manner. In a retrospective, single-center study on patients diagnosed with PAH, Shah et al evaluated patients’ adherence to phosphodiesterase-5 inhibitor therapy within an integrated pharmacy practice model at an outpatient pulmonary clinic at an academic institution. The pulmonary clinic collaborated with a specialty pharmacy and incorporated a clinical pharmacist and pharmacy technician as part of the healthcare team. In the integrated care model, clinical pharmacists provided comprehensive medication management, patient education, ongoing treatment monitoring, and assistance with transitions of care. Shah et al found that for the 131 patients in their analysis, the mean proportion days covered (PDC) was 96% (SD= 0.092), which is above the study’s defined adherence threshold of 80% PDC.^11^ The responsibilities of the pharmacist in this model are similar to the type of interventions performed by the pharmacist in our study. The models slightly differ as our study used collaborative practice agreement between the physician and clinical pharmacist rather than specialty pharmacy. The pharmacist’s role differs between a specialty pharmacy model and a collaborative care practice model. In a collaborative care practice model, such as our study, the clinical pharmacist played a role in managing dosing titration and closely monitoring patients to ensure they tolerated the titration well. Collaborative care practice pharmacists can also work closely with physicians regarding recommendations and send prescriptions directly to the pharmacy. This suggests that there could be a benefit to integrating pharmacists in the outpatient PH clinic as they can improve dose titration tolerability, medication access, and higher touch points of care in patients with PAH.

Clinical pharmacists can play an important role as part of the interdisciplinary team in the outpatient PH clinic setting using a collaborative care practice model approach. They can manage titration of medication and adverse events in complex patients who may have more severe PH and ensure prior authorizations are approved in a timely manner. Further research is warranted to measure how clinical pharmacists’ interventions lead to better outcomes, such as medication adherence, patient persistence rates, and time and money saved as part of the interdisciplinary team. Future research can also closely measure titration strategies and duration by a clinical pharmacist compared to the standard of care without a clinical pharmacist on the interdisciplinary medical team and assess how that impacts patient outcomes.

### Limitations

Limitations include the study design as a retrospective chart review, a small sample size from a single-center institution, descriptive data, and utilization of some baseline data that may not have aligned with the index pharmacy visit date. While the results of our study may not be able to be generalized to all patients diagnosed with PAH at other centers, we hope the interventions of clinical pharmacists at our center can help inform program development in other institutions.

### Conclusions

Clinical pharmacists can play an important role in closely monitoring patients during the medication titration phase and ensure prior authorizations are approved in a timely manner as part of the interdisciplinary team in the outpatient PH clinic setting using a collaborative practice model approach.

## Data Availability

Data is encrypted in an Excel sheet within the UCSF server.

## Acknowledgements

The authors would like to thank Sylvia Choi, for helping with chart reviews; Jennifer Cocohoba, PharmD, for helping to review the manuscript; and the UCSF IT team for generating an initial list of patients to begin chart reviews.

